# Reduced arousal during reward anticipation in unmedicated depressed patients

**DOI:** 10.1101/2020.03.03.20030478

**Authors:** Max Schneider, Immanuel G. Elbau, Taechawidd Nantawisarakul, Dorothee Pöhlchen, Tanja Brückl, BeCOME working group, Michael Czisch, Philipp G. Saemann, Michael D. Lee, Elisabeth B. Binder, Victor I. Spoormaker

## Abstract

Depression is a debilitating disorder with high prevalence and socioeconomic cost, but the central processes that are altered during depressive states remain largely elusive. Here, we build on recent findings in macaques that indicate a direct causal relationship between pupil dilation and anterior cingulate cortex mediated arousal during anticipation of reward. Using pupillometry and concurrent fMRI in a sample of unmedicated participants diagnosed with major depression and healthy controls, we observed reduced pupil dilation during reward anticipation in depressed participants with acute symptomatology. We further observed that individual differences in arousal during reward anticipation track the load and impact of depressive symptoms, a correlation that we replicated in a second sample of unmedicated depressed participants. Moreover, these group differences and correlations were mirrored at the neural level. The upregulation and maintenance of arousal during reward anticipation is a translational and well-traceable process that could prove a promising gateway to a physiologically informed patient stratification.

## Introduction

Major depressive disorder (MDD) is a mental disorder with a high prevalence and is estimated to be the top contributor to non-fatal health loss globally (World Health Organization, 2017). Experimental work has revealed that disturbances in the positive valence domain – as assessed with reward tasks – may be central to depressive symptomatology (Nestler and Carlezon, 2006; Kupfer *et al*., 2012; Russo and Nestler, 2013; Whitton *et al*., 2015).

The reward system comprises of various sub-processes that serve different functions within the anticipation, approach and consumption of rewarding stimuli. Most studied in the context of depression to date has been the processes of reward prediction and prediction error signaling. This followed the hypothesis that the common finding of anhedonia in depression mirrors deficits in predicting the value of an anticipated stimulus. Several studies have indeed observed group differences in the striatum during reward anticipation, when the task contained a learning component (Kumar *et al*., 2008; Gradin *et al*., 2011; Kumar *et al*., 2018b). This was not the case in a non-learning task: Rutledge *et al*. (2017) showed that individuals with depression exhibit reward prediction and prediction error signals in the ventral striatum similar to controls. Based on this, it was proposed that evidence for attenuated prediction error signaling in depression could mirror downstream effects more closely related to aberrant behavior. One such downstream subprocess is the upregulation and sustainment of arousal after reward prediction. Rudebeck et al. (Rudebeck *et al*., 2014) used pupillometry to track sustained arousal to an expected reward in macaque monkeys, and revealed that a lesion in the subgenual anterior cingulate cortex (in a few cases extending to the dorsal anterior cingulate) impaired sustained arousal in a reward delay task. Anatomical evidence indicates direct functional connections from the dorsal anterior cingulate and medial prefrontal cortex to the locus coeruleus (the main noradrenergic output center in the brain) in macaque monkeys (Aston-Jones and Cohen, 2005); this provides an anatomical pathway through which reward-induced salience could lead to physiological arousal.

In our previous pupillometry / functional magnetic resonance imaging (fMRI) study (Schneider *et al*., 2018), we translated these findings to healthy subjects by employing a well validated reward anticipation task (Knutson *et al*., 2001). We observed that the change in pupil size (*pupil dilation*) during reward anticipation was associated with activity in the dorsal anterior cingulate and bilateral insula, i.e., the salience network. This upregulation of arousal during reward anticipation likely facilitates reward approaching behaviors, which is in line with our previous observation that pupil dilation is correlated with (reward-associated) response times to a target stimulus (Schneider *et al*., 2018).

The goal of this study was to examine whether the ability to upregulate and sustain arousal during reward anticipation is different in a group of unmedicated, depressed participants in comparison to healthy controls. We used simultaneous pupillometry and fMRI measurements during the above-mentioned reward anticipation task in a sample of 41 unmedicated participants with major depression, including subthreshold major depression, and 25 control participants. Our hypotheses were that depressed participants would have reduced arousal, manifest in reduced pupil dilation, during reward anticipation compared to healthy controls, and that this would correlate with depressive symptom load and more specifically, anhedonia. Anhedonia was selected, because of the intrinsic conceptual relationship between reward related processes probed with our paradigm and anhedonia (Heshmati and Russo, 2015). Moreover, we hypothesized reduced activity in regions of the salience network during reward anticipation in depressed participants – the dorsal anterior cingulate and bilateral insula. In an exploratory analysis, we additionally addressed two core regions of the anti-correlated default mode network (posterior cingulate and medial prefrontal cortex), to examine if reduced *de*activation of this core regions was present in depressed participants.

## Materials and Methods

### Subjects

One hundred sixty-one subjects were recruited as part of the BeCOME study (“Biological Classification of Mental Disorders”; Brückl et al., submitted; registered on ClinicalTrials.gov: NCT03984084) conducted at the Max Planck Institute of Psychiatry (MPIP). All subjects underwent a general medical interview and an anatomical MRI screening to rule out present / past neurological disorders or any structural brain abnormalities. In addition, all subjects participated in an intensive psychometric assessment, which involved the computer-assisted Munich version of the Composite International Diagnostic Interview (DIA-X/M-CIDI - (Wittchen *et al*., 1998), which was adapted to the BeCOME study for the assessment of current (past two weeks) symptoms of depression and anxiety. Moreover, a battery of psychometric questionnaires was included in the study (Brückl *et al*., submitted manuscript), including the Beck Depression Inventory II (Kuhner *et al*., 2007). See ‘supplemental methods’ for more details.’

After screening for exclusion criteria and QC for pupillometry data, 41 unmedicated depressed patients fulfilled the inclusion criteria (current DSM-IV criteria for threshold or subthreshold MDE within the last 12 months). Subthreshold depression was defined as falling short of either the symptom criterion (four instead of the mandatory five depression symptoms were reported) or the impairment criterion (symptoms were present but did not cause clinically significant impairment). Within this group of N=41 was a group of 23 patients who reported five or more symptoms in the last 2 weeks (mean = 13.4, standard deviation [SD] = 6.1). The other 18 patients fulfilled current or sub-threshold MDE criteria, i.e. within the last 12 months, but not during the last 2 weeks (less than five symptoms in the last 2 weeks, mean = 1.1, SD = 1.3). We therefore used both the whole group of depressed participants (N=41) and the depressed participants with acute symptomatology (N=23) in our comparisons with healthy control participants. The healthy control participants consisted of twenty-five individuals who did not report any lifetime diagnosis. Age range, mean and variation as well as gender were similar in the groups (healthy control participants [N=25]; age range: 20 – 61 years, mean age = 32.1, SD = 10.3, 12 female; depressed participants [N=41] range: 19 – 64 years, mean age = 35.9, SD = 13.4, 27 female). See ‘supplemental methods’ for more details on inclusion and exclusion criteria, as well as QC of pupillometry data).

The study protocol was in accordance with the Declaration of Helsinki and approved by a local ethics committee. All participants provided their written informed consent after the study protocol had been fully explained and were reimbursed for their participation.

### Paradigm

Subjects performed a reward anticipation task (adapted from Knutson *et al*., 2001) inside the MR scanner, while pupil size was recorded using an MR-compatible eye tracker. The task utilized the same three conditions as in the original task: a potentially rewarding response condition (referred to as reward stimulus), a neutral response condition (neutral stimulus) and a control condition with no response-requirement (non-response control stimulus). Condition cues consisted of isoluminant gabor patch stimuli with different stripe orientations that were presented for 6 s respectively. In both response conditions, a light flash occurred after the 6 s anticipation time window, requiring a quick button press response to either obtain a monetary reward (1 €) or just feedback (a green checkmark symbol). If the response was too slow, a red cross appeared instead. The response conditions were followed by a number indicating the current cumulative total win (e.g. “3 €” following the third successful monetary reward trial). An adaptive algorithm ensured that participants would succeed on approximately 50% of reward trials across the session. During control trials, no flash was presented and no response was required. For a more detailed description of the experiment, see our previous publication (Schneider *et al*., 2018). After receiving task instructions, subjects completed an identical, two-minute training version of the task outside the scanner.

### Behavioral Data

To compare RTs between the reward and neutral stimuli and between the groups, we computed the median RTs across respective trials for each individual.

### Psychometric Data

Depression and anhedonia severity were determined using the data from the M-CIDI. We considered the following aspects/items within the E section of the M-CIDI: 1) number of depressive symptoms in the last 2 weeks (E51A1), 2) extent to which symptoms caused impairment in daily life functioning within the last 4 weeks (E53), 3) acuteness of depression (Items E2REC & E43REC), and 4) a mean score of the anhedonia-specific items loss of pleasure (E13 & E27), loss of appetite (CE15) and loss of sexual interest (CE26). These scores were later used to explore correlations between depressive / anhedonia symptomatology and pupil readouts / BOLD activity.

### Pupillometry

Pupil size of the subject’s right eye was recorded at a sampling rate of 250 Hz using an MR-compatible eye tracker (EyeLink 1000 Plus, SR Research), which was placed at the end of the scanner bore and below the presentation monitor. For a more detailed description of the post processing of the pupil data (including correction for gaze shifts and eye blinks), see ‘supplemental methods’ and (Schneider *et al*., 2018)

We averaged pupil size and dilation (the first derivative of size) over the whole stimulus duration (0-6 s) to avoid multiple per-second comparisons. For group comparisons (see Statistical Analyses), we used mean pupil dilation for each of the three stimuli [reward, neutral and no-respone control]. For correlative analyses, we again used the mean scores of the whole stimulus duration for pupil dilation, but we computed differential scores both for reward versus neutral (both conditions requiring a response) and for reward versus non-response control condition since the relative change was the primary variable of interest in these analyses.

To investigate the relationship between reward anticipation-related arousal and depressive symptomatology / anhedonia, we computed the correlation coefficients between pupil size and dilation and three psychometric scores of interest: the M-CIDI-based number and impact of depressive symptoms (within the last 2 weeks) and the M-CIDI-based anhedonia score. This analysis was first performed including all study participants (i.e., healthy controls and depressed participants). To rule out that potential correlations were driven by differences between healthy control and depressed participants, we repeated this analysis including depressed participants only. For correlations between pupil size and dilation and RT, we computed correlation coefficients across participant groups between pupil size and dilation and the median RTs for the two stimuli that required a response [reward and neutral stimulus].

### fMRI

Participants were scanned in a 3 Tesla MRI Scanner (MR750, GE, Milwaukee, USA) using a 32-channel head coil, covering 40 slices (AC-PC-orientation, 96 × 96 matrix, 2.5 mm slice thickness, 0.5 mm slice gap, resulting voxel size 2 × 2 × 3 mm^3^, echo planar imaging [EPI], TR 2.5 s, TE 30 ms, acceleration factor 2). Preprocessing and denoising was kept in accordance with our methodological validation study(Schneider *et al*., 2018).

The design matrices in SPM8 (https://www.fil.ion.ucl.ac.uk/spm/software/spm8/) involved one regressor per stimulus type. The following stimulus contrasts were obtained at the first level: “reward > non-response control stimulus” ([-1 0 +1]). “both response stimuli > non-response control stimulus” ([−2 +1 +1]) and the +1 contrast of the monetary reward stimulus. To test for differences between the control and depression group, two sample t-tests were conducted using the first level contrast images.

As we had strong hypotheses about regions of interest in the salience network, we extracted the individual betas (contrast reward > non-response control) from a 6 mm sphere around the peak voxel in the dACC, left insula and right insula. From the negative contrast we extracted the betas from two main default mode network regions: the medial prefrontal cortex and posterior cingulate (see Supplementary Table S1 for peak voxel coordinates). We additionally extracted the betas from the left and right ventral striatum.

### Statistical analyses

Group comparison analyses were Bayesian analyses as implemented in the software package JASP 0.9.2.0. (https://jasp-stats.org/). Group comparisons were performed with Bayesian repeated measures ANOVAs with group as a between-subjects factor, stimulus as a within-subjects factor, and age and gender as covariates (see ‘supplemental methods’ for further details on Bayesian inference and Bayes Factors).

Furthermore, in a Bayesian correlation analysis of pupil size with the total number of depressive symptoms, the observed data were modeled as samples from a multivariate Gaussian distribution (Lee and Wagenmakers, 2013; Matzke *et al*., 2017). Wide uniform priors were set for the mean of the (z-transformed) pupil dilation (−2,2) and of depressive symptoms (0,20), as well as for their standard deviations (0,2) and (0,10), respectively. The prior for the Pearson correlation was uniform as well (−1,1). The measurement error was incorporated into the model by adding one additional modeling step: the observed data were modeled as samples from true scores coming from a normal distribution with a given measurement error. The test-retest correlation was used to provide an estimate for the standard error of measurement: measurement SD × sqrt (1 – R_TEST-RETEST_), see ‘supplemental methods’ for further details on the estimation of test-retest correlations.This analysis was implemented in Matlab 2018a (www.mathworks.com) and JAGS 4.3.0 through the Matlab trinity interface https://github.com/joachimvandekerckhove/trinity.

Finally, group comparisons with fMRI were analyzed within the Bayesian framework implemented in SPM12 (https://www.fil.ion.ucl.ac.uk/spm/software/spm12/). Here a minimum threshold was selected (effect size = 0.5, BF ∼ 20) that has been shown to be more conservative than p_FWE_.cluster < 0.05 but more sensitive than p_FWE_.voxel < 0.05 in the frequentist approach (Han and Park, 2018). This threshold was increased for the stimulus contrasts (effect size = 1.0, BF ∼ 1000) to prevent single clusters from merging into one large cluster.

### Data availability

Raw data were generated at the MPI of Psychiatry. Derived data supporting the findings of this study are available from the corresponding author on request.

## Results

### Reduced arousal during reward anticipation in depressed participants

Healthy controls exhibited a strong pupil dilation in response to the reward stimulus, a moderate pupil dilation in response to the neutral stimulus, and a constriction of the pupil during the non-response control stimulus (**Error! Reference source not found**.A left and right panels).

For the comparison the whole depressed participant group versus healthy controls (Fig. 2B and 2C), we found no evidence for the models comprising group effects or interactions (Supplementary Fig. S1 and Supplementary Tables S2-3), although it provided overwhelming evidence for models including the stimulus effect (P[Model given data] = 0.999 for the 12 models containing the stimulus effect versus P[Model given data] = 0.001 for the aight models without the stimulus effect).

**Figure 1.**
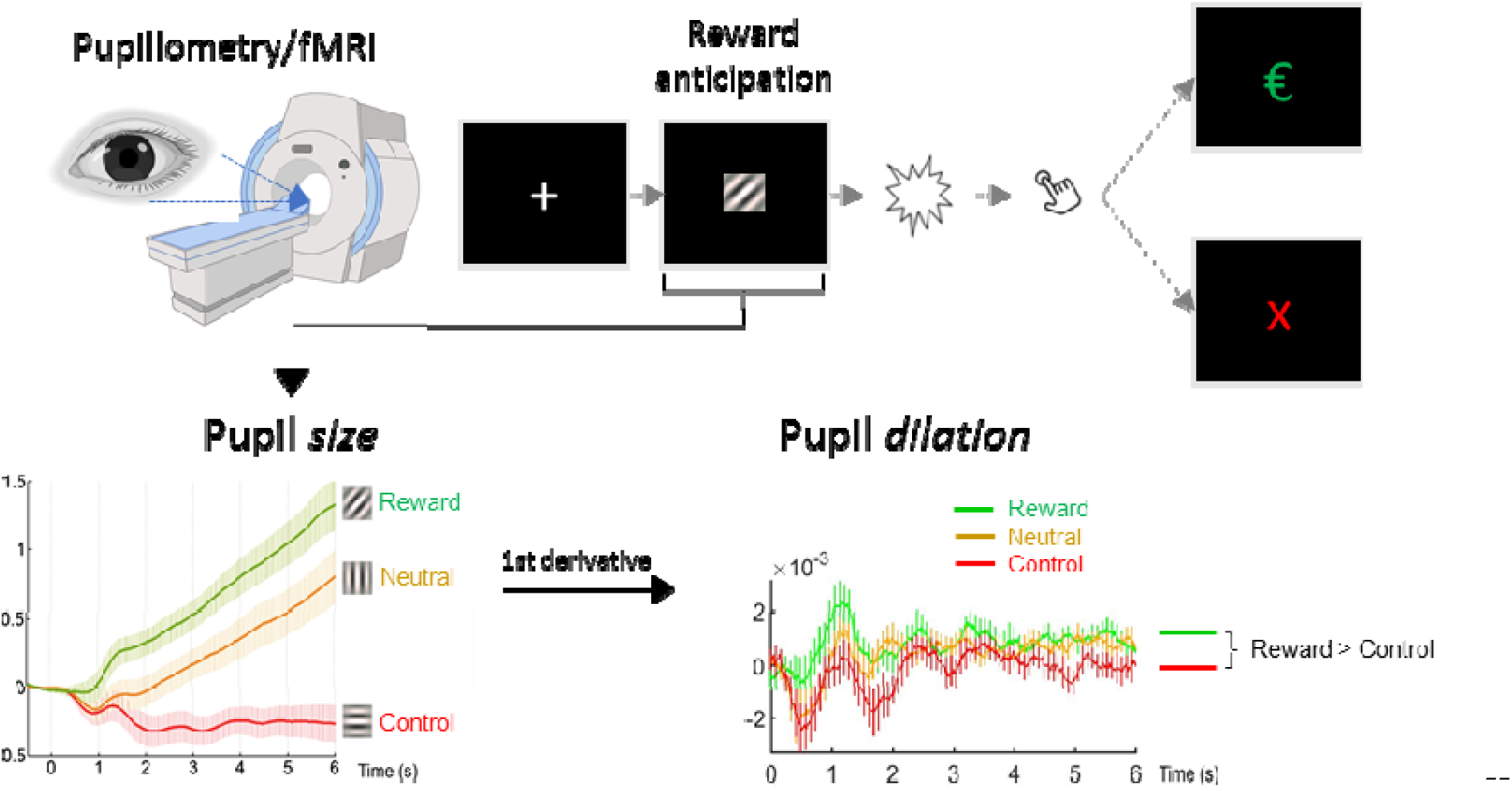
Graphic depiction of the reward anticipation task. Forty-one depressed participants and 25 healthy subjects performed an adapted version of the monetary incentive delay task by Knutson et al. (2001), with simultaneous pupillometry and fMRI. Three isoluminant gabor patch stimuli with different stripe orientations were presented for 6 s: after two of the stimuli, a light flash occurred that required a quick button press response to either obtain a monetary reward (reward stimulus) or a green checkmark symbol (neutral stimulus). There was no light flash and button press required for the third stimulus (non-response control stimulus). Pupil size was analyzed for the 6 s reward anticipation period; the increase or decrease of pupil size over time (first derivative) was used to quantify pupil dilation. Pupil dilation was averaged over the full anticipation period to obtain a robust score per stimulus and for the difference between the reward and non-response control stimulus. Values on the X-axis reflect time in seconds, on the Y-axes pupil size/dilation in z-transformed units.

**Figure 2.**
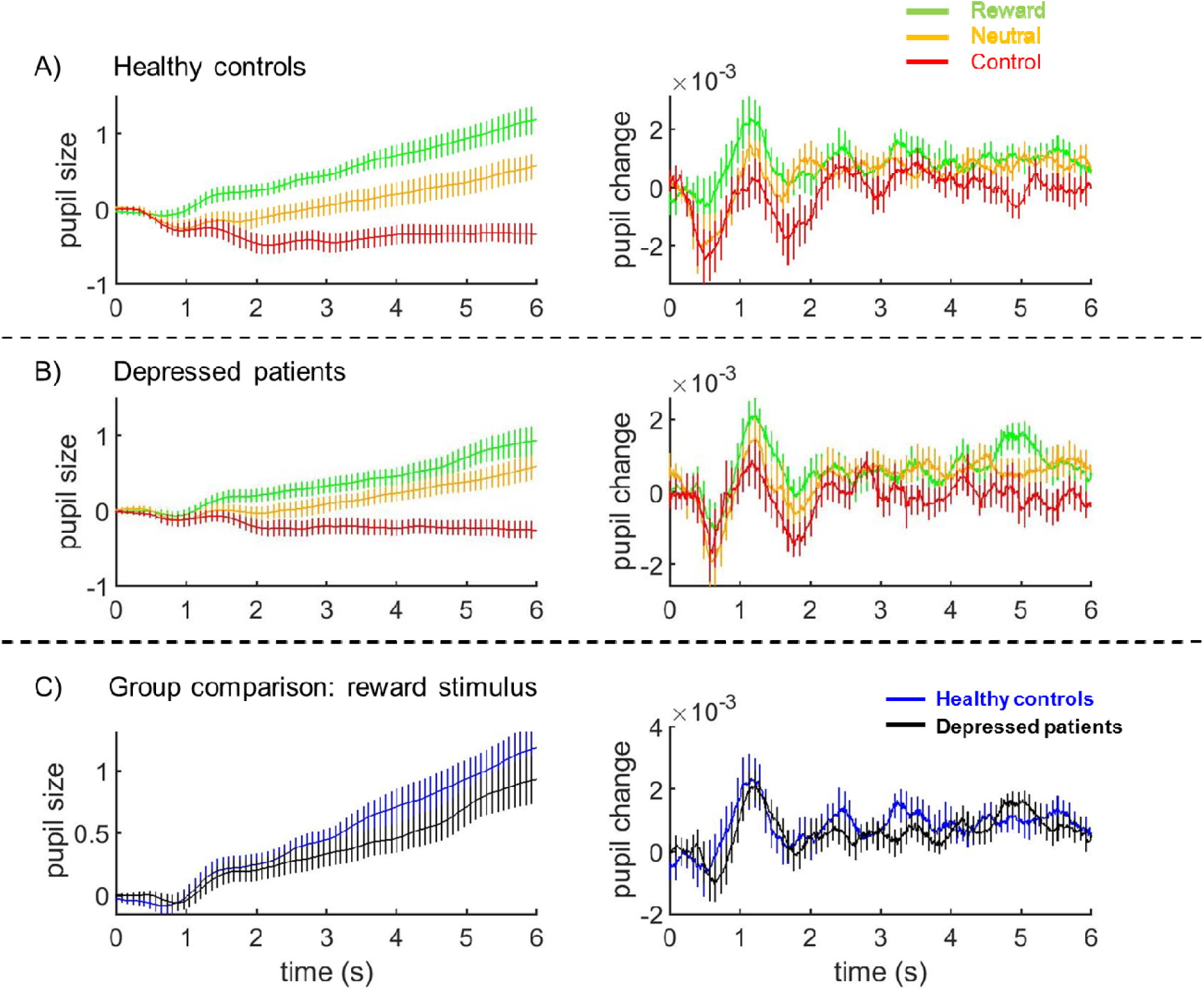
Pupil size and dilation during stimulus presentation per group. Pupil size (left panels) and pupil dilation (right panels) during presentation of the three stimuli in healthy control subjects (A) and depressed participants (B), averaged across all subjects and trials. C) Average pupil size and pupil change/dilation during presentation of the reward stimulus. Pupil values are z-transformed, vertical bars represent 95% confidence intervals.

For the comparison of the group of depressed participants with acute symptomatology (5 or more symptoms within the last 2 weeks) versus healthy controls (see Fig. S2 for descriptive plots), the same Bayesian repeated measures ANOVA yielded convincing evidence for the models comprising group effects or interactions (P[Model given data] = 0.887 for the four models containing the group × stimulus interaction versus 0.113 for the 16 models without this interaction, see Supplementary Fig. S2-3 and Supplementary Tables S4-5).

### Pupil dilation during reward anticipation correlates with depressive symptoms in a continuous manner

To examine whether the physiological process deviations underlying depressive symptomatology were continuous, we evaluated whether pupil dilation and depressive symptoms are correlated in a ‘dose-dependent’ manner across groups. To this end, we correlated (mean) pupil dilation during anticipation of reward with the number of current depressive symptoms (past two weeks), the impact of depressive symptoms (i.e., degree of impairment caused by these symptoms) and the sum of anhedonia-specific symptoms across healthy controls and depressed participants. Anhedonia specific symptoms were selected because of the intrinsic conceptual relationship between reward related processes probed with our paradigm and anhedonia (Heshmati and Russo, 2015).

There was very strong evidence for negative correlations of pupil dilation during reward anticipation with both number and impact of depressive symptoms (r = −0.52 and −0.46, BF_10_ = 2363 and 284, respectively), and strong evidence for a negative correlation with anhedonia specifically (r = −0.39, BF_10_ = 23.2). Partial correlation analyses controlling for age and gender provided strong evidence for the same negative correlations; r-values fell between −0.41 and −0.26.

Since these correlations could be confounded by group differences in pupil dilation and M-CIDI-scores, we re-ran them in the depressive participant group alone (N=41) (Fig. 3). There was again strong evidence for a negative correlation between pupil dilation and both number and impact of depressive symptoms (r = −0.53 and −0.45, BF_10_ = 80.9 and 13.2, respectively), but not for a correlation with anhedonia (r = −0.33, BF_10_ = 1.8) within the depressive participant group alone (see Fig. 2).

**Figure 3.**
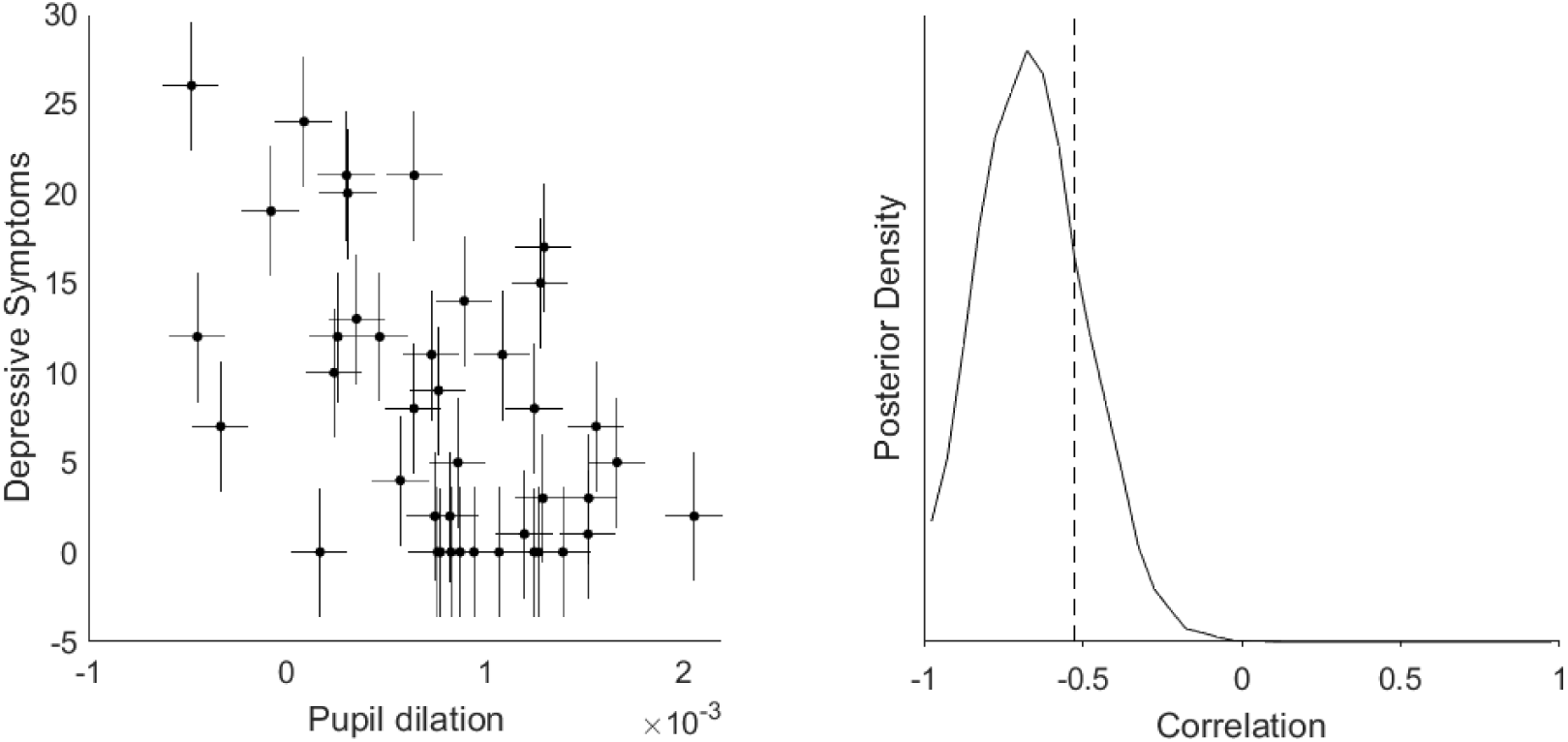
Correlation between pupil dilation and number of current symptoms with measurement uncertainty in depressed participants. Estimation of the correlation between the number of current depressive symptoms and pupil dilation to the reward stimulus with horizontal and vertical error bars representing the measurement uncertainty of the respective readouts (left panel). These measurement errors were incorporated into a Bayesian model that estimated the true correlations from observations sampled from a multivariate Gaussian distribution. The posterior distribution (right panel) provided very strong evidence that the true correlation is more negative than −0.3 (the dashed line represents the actual Pearson correlation value); the BF_10_ for r < −0.3 = 42.2).

To assess the specificity of these correlations, we additionally correlated pupil dilation in response to the three stimuli directly (as opposed to using the difference in dilation between the reward and non-response control stimulus) with the impact and number of depressive symptoms, as well as anhedonia. There was no evidence for any correlation of pupil dilation to the neutral stimulus and the non-response control stimulus, all r-values fell between −0.25 and 0.22, with BF_10_ values ranging from 1/2 to 1/5 (which represents anecdotal to moderate evidence in favor of the null hypothesis, i.e., no correlation). The evidence for correlations of pupil dilation to the reward stimulus matched the findings described above for the difference scores: very strong evidence for a negative correlation with number of depressive symptoms (r = −0.55, BF_10_ = 151.6), strong evidence for a negative correlation with impact of depressive symptoms (r = −0.49, BF_10_ = 33.2), and again only some evidence for a negative correlation with anhedonia (r = −0.38, BF_10_ = 3.3).

### Effects were specific to reward anticipation, not motor preparation

To examine whether the group differences and correlation were not merely due to the motor response (and/or the motor preparation associated with it), we re-ran the Bayesian repeated measures ANOVA of the acutely depressed participant group versus healthy controls, using the difference score of reward versus neutral stimulus, both of which require a motor response. This again yielded convincing evidence for the models comprising group effects or interactions (P[Model given data] = 0.646 for the four models containing the group × stimulus interaction We also re-ran the correlation analysis with the difference score of reward versus neutral stimulus (Fig. S4) and could replicate the correlations that we reported above: There was moderate evidence for a negative correlation between pupil dilation and both number and impact of depressive symptoms (r = −0.37 and −0.37, BF_10_ = 3.2 and 3.1, respectively), as well as for a correlation with anhedonia (r = −0.40, BF_10_ = 5.1) within the depressive participant group. The somewhat weaker effect might point towards a partial involvement of motor-preparatory processes that are correlated with increase in arousal during reward anticipation.

### Pupil dilation during reward anticipation is correlated with response time

We found very strong evidence for a negative correlation between pupil dilation to the reward stimulus and the median response time (RT) to the same stimulus across control subjects and depressed patients, r = −0.48, BF_10_ = 581.7 (Supplementary Fig. S4). Similarly, a negative correlation existed between pupil dilation to the neutral stimulus and the median RT to the same stimulus, r = −0.50, BF_10_ = 1349.1. This indicates that pupil dilation during reward anticipation tracks a process that has functional relevance.

### FMRI analyses provide evidence for associations between the BOLD correlates of reward anticipatory arousal in the salience network and depression symptom load and impact

The stimulus contrast reward > non-response control (full stimulus durations) revealed very strong evidence for activity in the anterior and middle cingulate gyrus, supplementary motor area, inferior/middle occipital cortex, insula, thalamus, caudate nucleus and ventral striatum, brainstem and cerebellum (Fig. 4A). The reverse contrast revealed bilateral clusters of activation in posterior cingulate gyrus, precuneus, medial frontal regions, middle occipital gyrus, angular gyrus and temporal regions.

**Figure 4.**
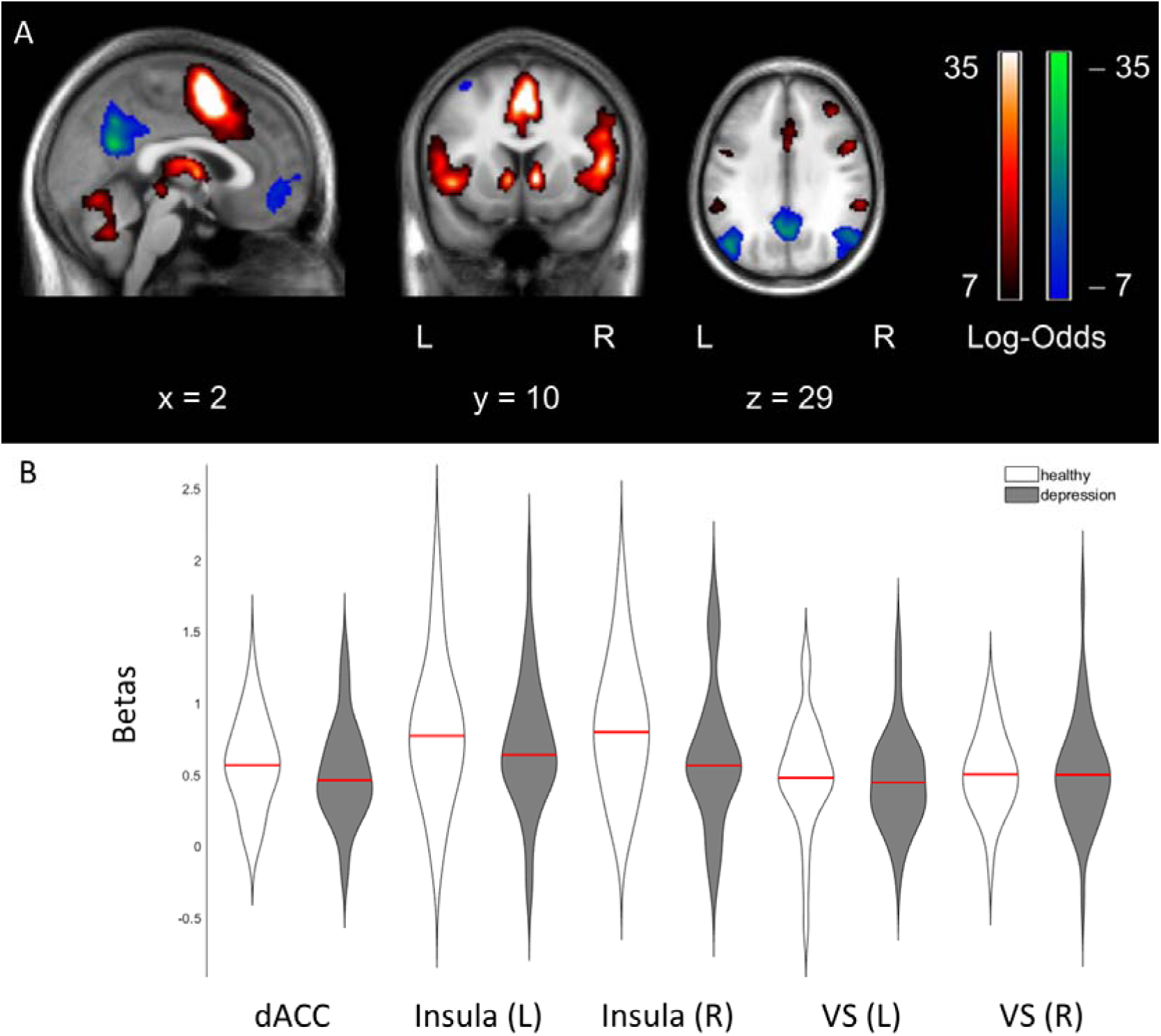
Statistical parametric maps of reward anticipation across participants and beta distributions per group. A) Neural correlates of a contrast representing reward > non-response control (hot colors) and its reverse contrast (cold colors) across participants, using a threshold with an effect size of 1.0 and BF_10_ = 1000. B) Violin plots for the regions of interest that depict the distribution of beta-values for the healthy controls (white) and depressed participants (grey) separately, with red lines depicting the group means. dACC, dorsal anterior cingulate cortex; L, left; R, right; VS, ventral striatum.

Since we had strong expectations about continuous effects and regions of interest in the salience network, we extracted the individual betas (contrast reward > non-response control) from a 6 mm sphere around the peak voxel in the dorsal anterior cingulate cortex (dACC) and left insula and right insula. In similar vein, we additionally extracted the beta estimates from the left and right ventral striatum given their relevance for reward anticipation (see Fig. 4B), and the medial prefrontal cortex and posterior cingulate cortex (PCC) as central nodes from the default mode network showing robust task deactivation (Supplementary Table S1 and Supplementary Fig. S5).

As with our pupil dilation analysis, comparisons for the extracted ROIs between depressed participants and control participants provided anecdotal to moderate evidence for the absence of group differences for all regions (BF_10_-values ranged from 1/4 to 1/2) except for the right insula, for which the BF_10_=0.8, providing neither evidence in favor of nor against group differences. However, when we again compared the group of depressed participants with acute symptomatology (5 or more symptoms within the last 2 weeks) with the control participants, we observed moderate evidence for group differences in the right insula (BF_10_ = 4.7) and in the PCC (BF_10_ = 5.1), with no convincing evidence in favor or against group differences for the dACC, left insula and the mPFC (BF_10_-values ranged from 1/2 to 1).

Of the three salience network regions, the correlations with the three clinical variables (symptom count, symptom impact and anhedonia, respectively) were all weakly negative within the depressive participant group, with moderate evidence for a negative correlation between right insula and number of depressive symptoms, r = −0.42, BF_10_ = 6.4. These correlations were in the same direction as correlations between pupil dilation and clinical variables albeit weaker, which is likely due to a reduced measurement certainty of the regional beta-estimates from the fMRI analysis. When we modeled in the measurement uncertainty in the correlation analysis between dACC and impact of depressive symptoms, there was moderate evidence for a correlation within the depressed participants, see Supplementary Fig. S6.

Of the default mode regions, all correlations with clinical variables were weakly positive, and there was only moderate to strong evidence for a correlation between the PCC and number of depressive symptoms, r = 0.36, BF_10_ = 13.5.

Finally, the two regions-of-interest that received moderate evidence for correlations with the number of depressive symptoms, also provided moderate evidence for a correlation with pupil dilation, r = 0.35, BF_10_ = 8.0, for the right insula and r = −0.35 for the PCC, BF_10_ = 7.0 across all participants. Within depressed participants, the correlation of PCC with pupil dilation was more strongly negative, r = −0.51, BF_10_ = 46.6, whereas there was no evidence any longer for (or against) a correlation of right insula with pupil dilation, r = 0.29, BF_10_ = 0.9.

### Replication of the correlation between pupil dilation during reward anticipation and current depressive symptoms

Since we made the cut-off for our initial analyses (September 2018), an additional 39 patients who experienced an MDE were enrolled in the study by June 2019. We did not observe evidence for differences in pupil dilation between participants who had an MDE in the last 12 months or before that (‘lifetime’), see Supplementary Fig. S7, and instead noted moderate evidence against between group differences, with BF_10_ = 1/3. Therefore, we included all 39 patients in a replication analysis to examine the evidence for correlations between pupil dilation (reward vs. non-response control) and number and impact of current depressive symptoms. There was again strong evidence for these correlations in the whole replication sample: r-values were −0.34 and −0.35, and BF_10_-values were 25.4 and 63.8, respectively (see Fig. 5). There was also moderate evidence for these correlations within the smaller group of participants who reported an MDE within the last 12 months (n = 29), r-values were −0.21 and −0.23, and BF_10_-values were 4.8 and 9.0, respectively.

**Figure 5.**
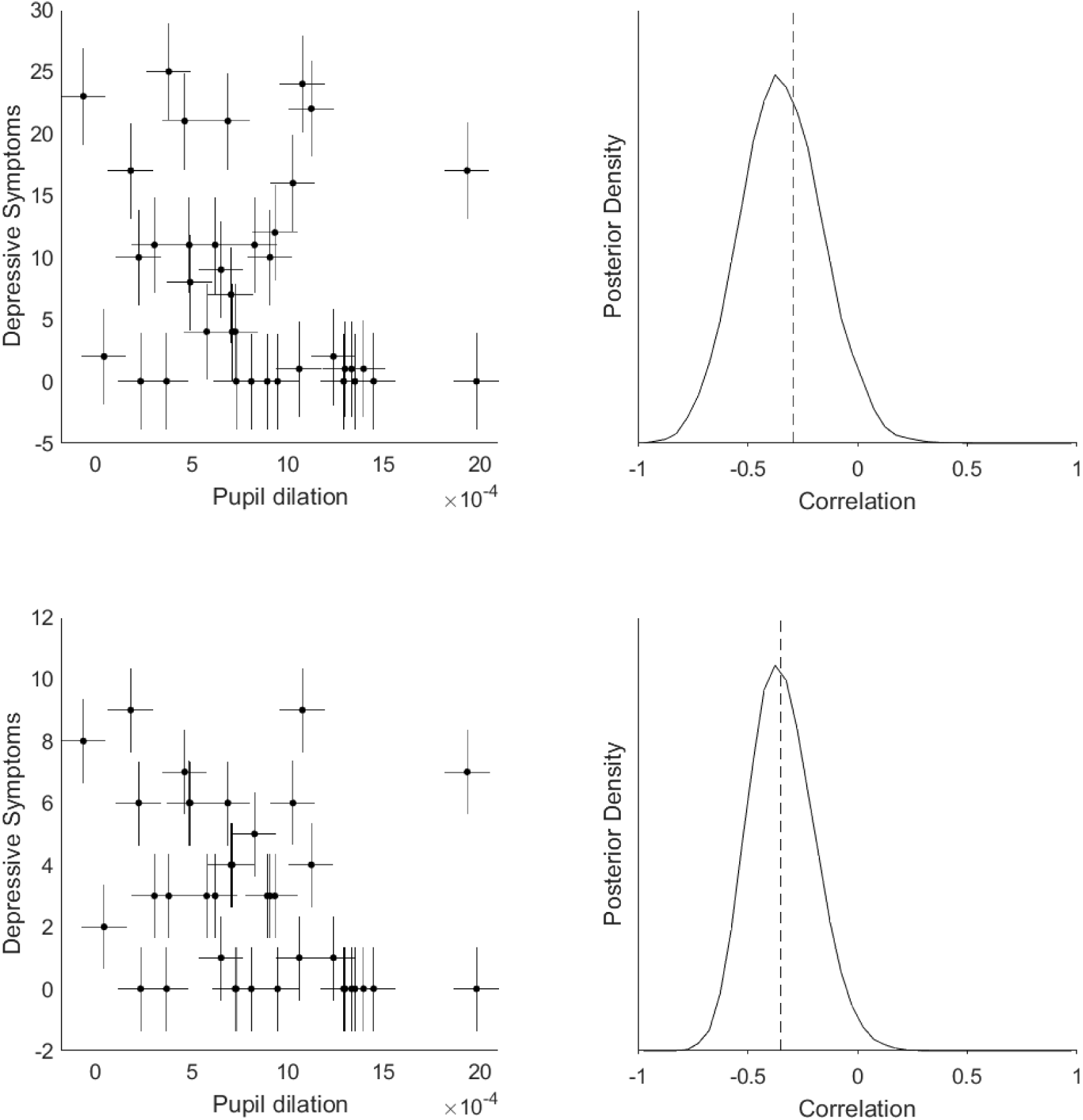
Replication of the correlation between pupil dilation and number of current symptoms in a second sample. Estimation of the correlation between the *number* of current depressive symptoms and pupil dilation to the reward stimulus in a replication sample (n=39), with horizontal and vertical error bars representing the measurement uncertainty of the respective readouts (upper left panel). The measurement error was estimated as in Figure 2, and they were again incorporated into a Bayesian model that estimated the true correlations from observations sampled from a multivariate Gaussian distribution. The posterior distribution (upper right panel) provided strong evidence that the true correlation negative (dashed line represents the actual Pearson correlation value of −0.35); the BF_10_ for r < 0 = 25.4). The lower two panels depict the same but for the *impact* of depressive symptoms, BF_10_ for r < 0 = 63.8.

## Discussion

We observed a robust negative correlation between reward anticipatory pupil dilation and depressive symptom load in two independent and similarly sized samples. This association existed both across healthy controls and participants labeled as having an MDE according to an extensive structured interview, as well as within the latter group only. This association translated to group differences between healthy controls and depressed participants, when we considere acute symptomatology (5 or more symptoms within the last 2 weeks). BOLD responses in the right insula (salience network) and the PCC (default mode network) correlated with pupil dilation and showed similar, but weaker correlation patterns with symptom load. In similar vein, task related activation in the insula and PCC showed moderate evidence for group differences between healthy controls and depressed participants with acute symptomatology. Moreover, we observed an inverse relationship between pupil dilation and response times, indicating that pupil dilation during reward anticipation has functional relevance. Taken together, these findings are in line with the notion that disturbances in the process of arousal regulation contribute to the phenotypic expression of depressive states.

### Interpretation of pupil responses as reward anticipatory salience/arousal transfer

We interpret the observed variance in pupil dilation as directly reflecting (state-) differences in anticipatory arousal during reward anticipation. This interpretation rests on experimental work in macaque monkeys that has demonstrated a direct causal link between the neuroanatomy of salience monitoring/arousal regulation and pupil dilation during reward anticipation (Rudebeck *et al*., 2014). We could previously translate these findings to humans by use of concurrent pupillometry/fMRI, where we found that reward anticipatory pupil dilation correlated with activity in the salience network (Schneider *et al*., 2018). In accordance with prevailing models on the involvement of the locus coeruleus (LC) in both pupil dilation and neural gain and optimal task performance (Aston-Jones and Cohen, 2005), we propose that after the initial reward prediction at stimulus onset, the transfer of reward prediction into action preparation might be the relevant process disturbed in participants with depressive symptoms. If LC activity is indeed lower, the neural gain signal that silences inactive regions and activates already active regions in order to prepare for the appropriate motor response, should become weaker, with reduced stimulus guided goal direction as a consequence. This hypothesis is also supported by strong evidence for a negative correlation between pupil dilation during reward anticipation and response times. Interestingly, a reduced drive to pursuit pleasurable (or necessary) activities – in German psychopathology “*Antriebsstörung*”, or “avolition” in English – is a common phenomenological feature that can be clinically observed in depressed patients. However, many self-rating questionnaires do not contain items to assess this phenomenological feature. As such, our results exemplify how objective measures of physiological processes could serve to stratify patients along dimensions that are not necessarily reflected in assessed symptoms.

Multiple other subprocesses are relevant to performing a reward task, such as reward prediction, decision-making and reward-consumption (Keren *et al*., 2018). While parallel contributions of these other subprocesses to depression pathology are likely (Starcke and Brand, 2012; Kumar *et al*., 2018a), it is unlikely that they underlie the observed pupil dilation differences within the reward anticipatory time window, since decision making related processes and reward consumption should manifest at different time points during the task. In line with this notion, a meta-analysis of six behavioral data-sets showed that depression specifically affected reward sensitivity rather than reward prediction or prediction errors (Huys *et al*., 2013). Regarding reward prediction error signaling, there is evidence for between group differences in the striatum when the task contains a learning component (Kumar *et al*., 2008; Gradin *et al*., 2011; Kumar *et al*., 2018b). However, this is not observed in a non-learning task (such as the one used in our study): Rutledge *et al*. (2017) showed that individuals with moderate depression exhibit reward prediction and prediction error signals in the ventral striatum that are similar to controls. Based on this, the authors concluded that depression does not affect (dopaminergic) reward prediction error signaling and that previous evidence for attenuated signaling could mirror downstream effects more closely related to aberrant behavior. The upregulation and sustainment of arousal after reward prediction is a downstream process more closely related to behavior – in our case it comprised a simple button press but in more complex tasks this could extend to approach behavior (Huys *et al*., 2016). It is of note that our pupillometry data during reward anticipation do not indicate differences in general arousal levels, but specifically in the upregulation of arousal for reward-associated motor preparation. General differences in arousal would have been reflected in differences in pupil sizes at the beginning of the anticipatory phase or to all stimuli, which was not the case. Finally, the continuous relationship between acute symptom load (past two weeks) and pupil dilation during reward anticipation, independent of previous MDE diagnosis, also hints towards a state-marker characteristic of reward anticipatory arousal. By contrast, aberrances in reward related learning signals have been observed in remitted depression, pointing towards the relevance of this process as a trait marker (Geugies *et al*., 2019). More evidence regarding such characteristics will be useful for potential applications as a treatment response tracking or drug target engagement measure.

### Similar but weaker associations between clinical variables and BOLD responses in regions of the salience network and DMN

Taken together, we observed similar, but weaker associations between depressive symptom load/impact and BOLD responses within the right insula and PCC during reward anticipation. These associations were paralleled by correlations between pupil dilation and BOLD responses in these regions, as in our previous methodological paper (Schneider *et al*., 2018). The weaker nature of these associations as compared to the correlations between pupil dilation and depressive symptom load directly, is likely due to the higher measurement uncertainty and generally lower test-retest reliability for single-subject regional fMRI beta-estimates (during reward tasks ∼0.5-0.6; (Plichta *et al*., 2012). In line with this notion, when we modeled the measurement uncertainty in the betas extracted from the dACC (reward > non-response control) in a correlation analysis with clinical variables, we actually observed moderate evidence for a correlation between impact of depressive symptoms and dACC. All in all, these results indicate that salience network regions correlate negatively with depressive symptom load, paralleling pupil dilation. This is interesting as it lends further evidence to the notion that fMRI measures of the salience network – such as resting state connectivity metrics – reflect a disease-relevant physiological process. This is in line with results from a recent meta-analysis that implicated disruption in the salience network in depression and trans-diagnostically across psychiatric disorders (McTeague *et al*., 2020). In contrast, the PCC – as part of the default mode network that deactivates in our task – shows the opposite pattern, which represents a failure to reduce default mode network activity during reward anticipation. This is in accordance with previous work that suggests differences in task-related default mode network activity in depressed participants (Sheline *et al*., 2009; Bartova *et al*., 2015).

### Advantages and limitations of pupillometry as a measurement tool for LC dependent NA activity

A compelling feature of pupillometry is that it provides indirect access to the central monoaminergic regulatory systems that are implicated in a wide variety of cognitive and emotional functions. This has been utilized in the past to characterize depressed population regarding differences in the physiological response to various tasks, such as working memory (Jones *et al*., 2010), cognitive control (Siegle *et al*., 2004; Jones *et al*., 2015) and emotional processing (Siegle *et al*., 2001; Silk *et al*., 2007; Steidtmann *et al*., 2010; Jones *et al*., 2011; Siegle *et al*., 2011; Burkhouse *et al*., 2015). Further, differences in the more direct autonomic regulation of the pupil have been studied in depressed populations, such as pupillary unrest as a measure of vigilance and spontaneous arousal fluctuations (Schumann *et al*., 2017), or the pupillary light reflex (Bar *et al*., 2004; Wang *et al*., 2014; Mestanikova *et al*., 2017; Sekaninova *et al*., 2019). Our works extends this body of work by highlighting the physiological process of arousal upregulation during reward anticipation in the pathophysiology of depression.

The finding of similar, but stronger patterns of associations between clinical variables and pupil dilation than between clinical variables and the fMRI correlate of pupil dilation might be due to the fact that pupillometry provides a more direct and potentially more sensitive tracking of relevant brainstem nuclei, like the LC, activity of which precedes pupil dilation by roughly 300 ms (Joshi *et al*., 2016a). However, as a surrogate marker for LC activity, the main origin of noradrenergic projections in the brain, pupillometry may not be informative for stages of the reward anticipation that are not LC dependent (such as reward prediction and consumption). Instead, pupil dilation should be expected to specifically track the LC dependent upregulation of arousal that occurs after the initial reward prediction and before the actual motor action. This is in line with our previous methodological work that examined the neural correlates of pupil dilation during reward, which specifically correlated to the dACC and bilateral insula (Schneider *et al*., 2018). In contrast, reward prediction at stimulus onset correlated with activity in ventral striatum, which is in line with the larger literature on reward prediction error (Garrison *et al*., 2013; Chase *et al*., 2015). Conversely, LC upregulation of arousal during the prediction of an aversive stimulus might be another process of interest.

Furthermore, although numerous studies provide strong evidence for a close relationship between pupil dilation and noradrenergic activity in rodents, primates and humans (Aston-Jones and Cohen, 2005; Murphy *et al*., 2014; Joshi *et al*., 2016b; Reimer *et al*., 2016), other neuromodulatory systems like the cholinergic and serotonergic system are known to influence pupil dilation as well (Larsen and Waters, 2018). For example, Reimer *et al*. (2016) found that in mice, spontaneous pupil fluctuations track both changes in noradrenergic and cholinergic activity.

Interestingly, they observed that fast pupil dilations typically occurring during rest and at the beginning of walking on a treadmill were closely linked to phasic noradrenergic activity, whereas longer-lasting dilations observed during continuous locomotion were accompanied by sustained cholinergic activity. Their finding seems to fit our interpretation of a noradrenaline-mediated upregulation of arousal that facilitates subsequent (goal-directed) behavior, however, we cannot rule out that pupil dilations observed in the present study also involve a cholinergic component. Another neurotransmitter that has been associated with changes in pupil size is serotonin: for example, serotonergic agonists like lysergic acid diethylamide (LSD) have been shown to cause a dilation of the pupil (Schmid *et al*., 2015), which has been suggested to result from interactions with LC neurons and subsequent release of noradrenaline (Yu *et al*., 2004; Einhauser *et al*., 2008; Larsen and Waters, 2018). Since patients included in the present study were unmedicated and had a similar baseline pupil size as healthy control subjects, we can rule out that the observed group differences in pupil dilation might reflect differences in anti-depressant medication. However, more studies will be needed to precisely disentangle noradrenergic, cholinergic and serotonergic influences on pupil size.

### Other methodological considerations

One further limitation of this study is the limited sample size of healthy controls in the original sample. This was in part due to the rigorous phenotyping of all subjects that included an extensive M-CIDI assessment, whereby a substantial portion of subjects that were recruited as healthy controls turned out to fulfill psychiatric diagnoses (see Methods). However, since we were primarily interested in continuous associations between differences in reward anticipatory arousal and depression symptom load, the role of healthy subjects was to span the variance in our study population and not to increase power for group comparisons. A further restraint on the overall sample size was a rigorous quality control of pupillometry data, which led to the exclusion of 48 participants. However, despite a limited sample size, we could strengthen the evidence for our observations reflecting true effects by reproducing our main findings in an independent sample. The overall slightly weaker effects for the correlation between pupil dilation and depressive symptom load in the replication sample, likely is due to inflated effect sizes in the initial sample or the “winner’s curse” (Costa-Font et al., 2013), with the true effects likely being more in the range of the replication analysis.

## Conclusion

In conclusion, our results suggest that disturbances in the physiological process of arousal upregulation in anticipation of predicted reward underlie in part the phenotypic expression of depression symptomatology. The ability to track this process directly in individual patients via pupil responses might be a promising gateway to a physiologically informed patient stratification and could inform novel interventions that target this specific process.

## Data Availability

Raw data were generated at the Max Planck Institute of Psychiatry. Derived data supporting the findings of this study are available from the corresponding author upon reasonable request.

## Acknowledgements

We would like to thank Stephanie Alam, Miriam El-Mahdi, Gertrud Ernst-Jansen, Carolin Haas, Karin Hofer, Elisabeth Kappelmann and Rebecca Meissner for their help with data collection, study management, the recruitment and screening of BeCOME participants, and Ines Eidner und Anna Hetzel for their assistance with MRI scanning. We thank Pamela Hathway for help in implementing the Reward Anticipation Task. Our special thanks go to all study participants for participation in the BeCOME study.

## Funding information

There were no external funding sources used for this study.

## Competing interests

The authors declare no competing interests

## Notes

### Competing Interest Statement

The authors have declared no competing interest.

### Funding Statement

No external funding received

